# Designing a children’s health exposomics study protocol: The CHILDREN_FIRST multi-country prospective cohort using multi-omics and personalized prevention approaches

**DOI:** 10.1101/2025.06.04.25329011

**Authors:** Corina Konstantinou, Georgia Soursou, Samuel Abimbola, Pantelis Charisiadis, Angelos Kyriacou, Theofano Modestou, Michalis Tornaritis, Charalambos Hadjigeorgiou, Agapios Agapiou, Efstathios A. Elia, George Milis, Alexis Kyriacou, Lygia Eleftheriou, Zoi Tsimtsiou, Pantelis Natsiavas, Or Duek, Idan Menashe, Nathalia Bilenko, Itamar Grotto, Enkeleint A. Mechili, Mònica Guxens, Costas A. Christophi, Constantinos Deltas, Konstantinos C. Makris

## Abstract

Non-communicable diseases (NCDs) account for ∼71% of all deaths globally, including 15 million premature deaths each year (deaths between 30-69 years of age). Instead of waiting until the disease manifestation, focusing on the origins of NCDs during childhood offers a critical window of disease prevention and control for effective interventions. The CHILDREN_FIRST study aims to investigate how the spatio-temporal evolution of the children’s exposome profiles in the Mediterranean region influences the early-life programming of chronic disease risk during the unique critical window of susceptibility in the primary school years (6-11 years of age). The study protocol adopts the human exposome framework integrated with a personalized prevention approach using multi-omics platforms and advanced machine learning algorithms implemented across five Mediterranean countries, namely Cyprus, Greece, Spain, Israel, and Albania. The cohort will consist of children enrolled in the first grade of primary school, who will undergo annual follow-up assessments until completion of primary education. During the annual assessments, children’s exposome parameters from the three main exposome domains will be evaluated using different assessment types i.e., biospecimen, sensors, questionnaires. Standardized human sample and data collection methods will be employed following harmonized standardized operating procedures. The reference model of Observational Medical Outcomes Partnership – Common Data Model part of the Observational Health Data Sciences and Informatics will be used to conduct federated data analysis. This CHILDREN_FIRST study protocol is a human exposome-based initiative to establish a long-term prospective cohort infrastructure for biomedical research on children’s health within the Mediterranean region. The cohort’s exposome-based findings will systematically feed into the evaluation and design of chronic disease prevention programs. Expected results would inform evidence-based policy making and the development of health interventions for reducing the risk of NCDs.

## 2 Introduction

Non-communicable diseases (NCDs) account for ∼71% of all deaths globally, including 15 million premature deaths each year (deaths between 30-69 years of age), thus, undermining workforce productivity and impeding economic growth (1). Among adolescents aged 10-24 years, NCDs are responsible for 86.4% of all years lived with disability (YLDs) and 38.8% of total deaths (2). Considering the global cancer burden, the leading risk factors with the highest number of age-standardized disability-adjusted life years (DALYs) remained the same between 2010 and 2019, with smoking, alcohol use and high body mass index (BMI) identified as the top contributors (3).

Global efforts adopt a life course approach to NCD prevention, acknowledging the value of addressing risk factors early in life (4) (5) (6). Instead of waiting until the manifestation of disease’s symptoms, the focus on the origins of NCDs during childhood offers a critical window of disease prevention and control for effective preventive interventions, rather than relying solely on prevention or treatment strategies later in adulthood. Children exhibit increased susceptibility to environmental exposures compared to adults because of their rapid development, differences in behaviors and metabolism, and indirect/passive exposures to multiple environmental stressors via their parents’ habits and lifestyle (7).

On the fetal programming of chronic disease, several birth-pregnancy cohorts have mostly collected baseline exposure data during the prenatal period and followed up during early life, while other cohorts continue extending longer their follow-ups into the primary school years and beyond. In particular, the age span of 6 to 11 years, being the first years of school life, carries some unique features. In effect, children enter primary school, spending more time in organized educational settings (schools), compared to when they were younger, and begin different hobbies, social interactions and friendships, while their learning experience gets steeper and often versatile in content and types (8). Based on the predicted obesity prevalence rates among 2-year-old children from the CHOICES simulation model, a steeper increase in obesity prevalence was estimated for ages between 5 to 11 years old (9) than younger age groups or older than 12 years of age. Moreover, longitudinal studies have shown that three out of six identified lung function risk trajectories - that contribute to 75% of chronic obstructive pulmonary disease (COPD) in adulthood - originate in childhood using data from seven-year-old children (n=2438) (10).

Holistic methodological frameworks, such as the human exposome concept and its exposomic tools are essential for supporting the implementation of children’s health policies (11); such frameworks would allow us to better understand the complex associations between environmental exposures and trajectories of healthy growth and development in children. The human exposome encompasses all environmental exposures from conception onwards, with the associated biological response (12,13). It was proposed in an attempt to elucidate the role of the environment in the development of chronic diseases (14) given that environmental factors may contribute up to 80-90% of NCD risks (15). In conjunction with the human exposome concept and its utility, the precision prevention approach within the personalized medicine paradigm emerges as a new medical model that seeks to characterize individual phenotypes and genotypes to determine disease predisposition and to deliver timely prevention (16). When applied to children’s populations, precision prevention would study differences in children’s genetic makeup, environmental exposures, and lifestyle factors. This approach not only provides knowledge about enhanced risk profiling and population stratification but also holds promise for informing more effective individualized interventions to improve child health outcomes (17).

There are several children’s exposome projects to date, with most of them enrolling mothers during pregnancy (18,19). Such cohort exposome studies collect repeated in time sample/data prenatally, but they often go by a single assessment point during the 6-11 age period. Some children’s exposome studies pool data from existing birth cohorts, limiting the uniformity of exposure and outcome measurements and they may typically vary in recruitment and follow-up timings, children’s age at the time of the examination, data completeness, and biospecimen measurement types and tools. Harmonization of data collection tools and procedures across diverse settings and cohorts that pool data from heterogeneous cohorts and their populations is a daunting task (20,21).

In response to these methodological challenges in children’s health, we designed a multi-country site exposomic longitudinal cohort children’s study, during a critical window of vulnerability, using *a priori* harmonized protocols, that would enable its future adaptation or replication to a different population and setting around the globe. As a case study, we present the CHILDREN_FIRST exposomics-based study protocol for the multi-country prospective cohort study focused on children in Mediterranean populations. This longitudinal study aims to investigate how the spatio-temporal evolution of the children’s exposome profiles in the Mediterranean region influences the early-life programming of chronic disease risk during the critical window of susceptibility present in the primary school years (6-11 years of age). The CHILDREN_FIRST study adopts the human exposome framework and its exposomic tools together with a personalized prevention approach using multi-omics platforms and advanced machine learning algorithms.

The specific objectives of the CHILDREN_FIRST exposomics-based prospective cohort study are to: **i)** characterize the spatio-temporal variability of the targeted/untargeted multi-exposomic profiles of primary school children in Cyprus, Greece, Spain, Albania, and Israel, using standardized sample/data collection, biomarker and multi-omics capabilities and machine learning data processing algorithms, **ii)** evaluate the effect of single and multiple environmental exposures on early-life biological programming linked to chronic disease risk, by applying validated biomarkers of exposure and effect, related to cardiometabolic, neurodevelopmental, and respiratory outcomes and their early precursors, such as inflammation and oxidative stress/damage, including multi-omics, meet in the middle capabilities, and **iii)** generate NCD risk prediction groups of children using precision prevention approaches that include the spatio-temporal profiling of both genetic and non-genetic stressors, such as environmental, social, behavioral, metabolic, and psychosocial risk factors and multi-omics platforms.

## 3 Methods and analysis

### 3.1 Study Design

CHILDREN_FIRST is a multi-site, longitudinal cohort study to be implemented across Cyprus, Greece, Spain, Albania, and Israel. Schools will be the main setting for recruitment, assessment and reporting back, by enrolling children in the first grade of primary school, who will undergo annual follow-up assessments until completion of primary education (Figure 1). During the annual assessments, exposome parameters from the three main exposome domains will be evaluated.

**Figure 1.**
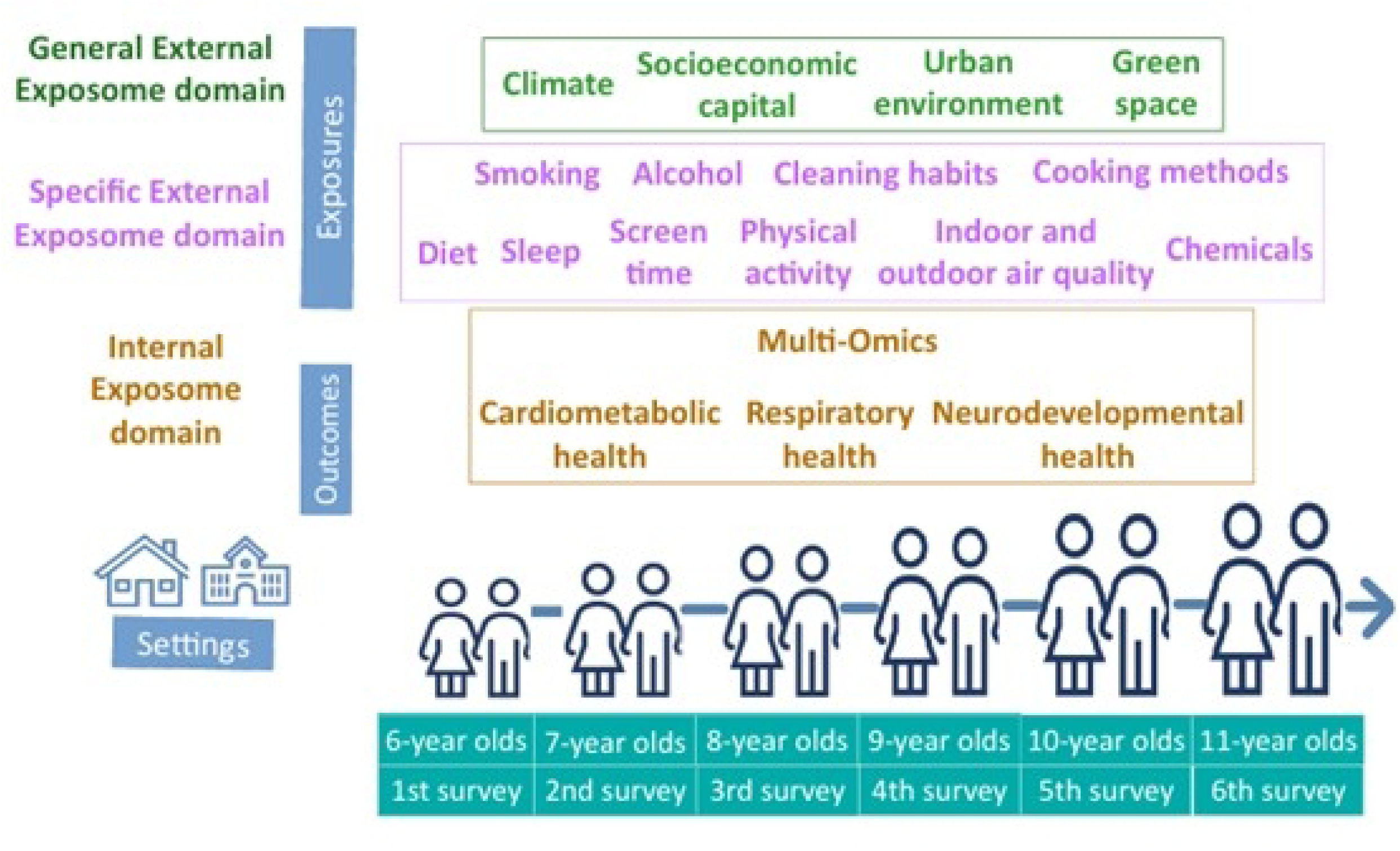
CHILDREN_FIRST study design: Annual children’s exposome assessments during primary school years (first grade to sixth grade).

### 3.2 Study Population

Randomly selected public primary schools will be the study’s main recruitment setting. All first-grade students attending the randomly selected schools and residing in the country for at least the past year will be eligible for inclusion. The number of participants recruited from each area will be proportionate to the population distribution of children, taking also into account the degree of urbanization and the administrative units of each country (e.g., districts, municipalities, etc.) (more about recruitment in Supplementary File 1).

### 3.3 Study procedures

Annual assessments of the children’s exposome will be conducted from the first through the sixth grade of primary school. Standardized human sample and data collection methods will be employed in all countries following standardized operating procedures (more about Assessment methods and Questionnaires list in Supplementary File 1). Table 1 outlines the variables to be measured (aggregated to a broad class variable), the corresponding assessment methods (e.g., electronic questionnaires, biospecimen) and their classification within the exposome framework (e.g. general external, specific external, or internal exposome domain).

**Table 1.** Overview of variables, assessment methods and exposome domains to be measured in the CHILDREN_FIRST prospective cohort study.

| Variable group | Assessment method | Exposome domain* |
| --- | --- | --- |
| Climate (ambient air temperature and relative humidity) | Sensors in schools | General external |
| Built environment (e.g. population density, building density, street connectivity) | GIS linkage to local spatial data, secondary data | General external |
| Neighborhood quality of life (e.g. health care access, life in the neighborhood and green urban spaces) | Questionnaire | General external |
| Socioeconomic capital (e.g. parental education and employment, household composition) | Questionnaire | General external |
| Outdoor air quality (e.g. CO<sub>2</sub>, PM, VOCs) | Sensors and passive samplers in schools | Specific external |
| Indoor air quality (e.g. CO<sub>2</sub>, PM, temperature, humidity, noise, UV, VOC) | Sensors and passive samplers in schools, questionnaire on school/classroom/home characteristics | Specific external |
| Residential and school environment (e.g. noise, traffic, ventilation, building materials, heating sources, cleaning practices, cooking methods, ALAN) | Questionnaire on school/classroom/home characteristics, satellite-based images of residences for outdoor ALAN | Specific external |
| Water contaminants (e.g. THMs, PFAS) | Tap water samples from houses and schools | Specific external |
| Child lifestyle e.g. physical activity, sedentary behavior, screen time, diet, sleep patterns, chronotype | Questionnaire | Specific external |
| Parental lifestyle (e.g. smoking, alcohol use, occupation) | Questionnaire | Specific external |
| Biomarkers of exposure (e.g. pesticides, metals, bisphenols, VOCs) | Urine and saliva samples<sup>^</sup> | Specific external |
| Background characteristics (e.g. age, sex) | Questionnaire | Internal |
| Systems biology (e.g. omics) | Urine and saliva samples | Internal |
| Oxidative stress/damage and inflammation | Urine samples | Internal |
| Iodine status | Urine samples | Internal |
| Circadian function (e.g. corticosteroids) | Urine and saliva samples | Internal |
| Liver and renal function | Urine samples | Internal |
| Medical history (e.g. health status, medicine use pregnancy/breastfeeding practices) | Questionnaire | Internal |
| Intrinsic properties (e.g. weight, height, waist circumference, birth weight) | Anthropometric measurements, questionnaire | Internal |
| Psychosocial health (e.g., stress, allostatic load, etc.) | Questionnaire | Internal |
| Neurodevelopmental disorders (e.g. ASD) | Questionnaire | Internal |
| Cardio-metabolic outcomes (e.g. BMI, blood pressure) | Anthropometrics, Sphygmomanometer | Internal |
| Respiratory health (e.g. asthma, wheezing) | Questionnaire | Internal |

\* Exposome domains based on (22).
^Noninvasive biospecimen matrices, such as hair, nails, are preferred instead of blood, because of the more frequent repeated measures nature of the CHILDREN\_FIRST exposomics protocol.
Abbreviations: GIS: Geographic Information System, CO<sub>2</sub>: Carbon dioxide, PM: Particulate matter, VOCs: Volatile organic compounds, ALAN: Artificial light at night, THMs: Trihalomethanes, PFAS: Per- and polyfluoroalkyl substances, ASD: Autism spectrum disorder, BMI: Body mass index

### 3.4 General External Domain

Climatic parameters, specifically air temperature and relative humidity, will be monitored using sensors installed in outdoor areas of the participating schools. Built environment characteristics, including population density, building density, and street connectivity, will be assessed through Geographic Information System (GIS) linkage to local spatial datasets and relevant secondary data sources. Standardized questions based on validated questionnaires will be used for the assessment of demographic characteristics and neighbourhood quality of life (e.g., life in the neighbourhood and green urban spaces) (23), health care access (24), sociodemographic variables (e.g., parents’ employment and education status, parental age, country of birth (grandparents, parents, and child), language spoken at home, years living in area, household income, number of people in household, family/cohabitation status, address, and contact info) (23,25,26).

### 3.5 Specific External Domain

#### 3.5.1 Air quality

Air quality parameters will be monitored in classrooms of participating schools (and potentially also in children’s residences) for a minimum of one school week, each year. Measurements will include PM_10_, PM_2.5_, air temperature, relative humidity, CO_2_, as well as volatile organic compounds (VOCs), such as benzene, toluene, ethylbenzene, xylene (BTEX), and chloroform.

#### 3.5.2 Water pollutants

All four trihalomethanes (THMs) species - chloroform (TCM), bromodichloromethane (BDCM), dibromochloromethane (DBCM), and bromoform (TBM) - will be measured in the collected tap water samples from the participating houses and schools. The THMs analysis will be conducted according to a previously published method (27). Per- and polyfluoroalkyl substances (PFAS) as well as microbiological parameters will also be assessed in tap water samples.

#### 3.5.3 Chemical contaminants

Numerous chemicals could be targeted here. Indicatively, we plan on using urinary biomarkers of exposure to pesticides, including two pesticide metabolites: 3-phenoxybenzoic acid (3-PBA), a biomarker of pyrethroid exposure, and 6-chloronicotinic acid (6-CN), a metabolite of neonicotinoid pesticides using a gas-chromatographic-tandem mass spectrometric (GC–MS/MS) method based upon modifications of two existing protocols, among others (28,29). Urinary concentrations of lead and cadmium will also be measured using the Centers for Disease Control and Prevention (CDC) multi-element ICP-DRC-MS method No. 3018.3 (30). Urine samples will be analyzed for free and total forms of bisphenols, such as bisphenol A (BPA) bisphenol F (BPF) or BPS and three chlorinated BPA derivatives: 3-chlorobisphenol A (ClBPA), 3,5-dichlorobisphenol A (3,5-Cl_2_BPA), and 3,3’-dichlorobisphenol A (3,3’-Cl_2_BPA), using a modified version of a previously published urinary BPA analysis protocol (31). A suite of VOCs will be assessed in saliva samples.

#### 3.5.4 School and home environment

Standardized questions based on validated questionnaires will be used for the assessment of residential environment and home exposures (23,26,32) and school building/classroom characteristics that potentially have an impact on indoor air quality (33). Indoor artificial light at night (ALAN) will be assessed using a subjective measure defined as the level of light in the bedroom during sleeping time (34). The response will be a four-digit Likert scale: a) total darkness, b) almost dark, c) dim light, and d) quite illuminated. For the evaluation of outdoor ALAN with calibrated satellite-based images of children’s residences with high-quality spatial resolution during the study period will be used.

#### 3.5.5 Parental and child lifestyle

Standardized questions based on validated questionnaires will be used for the assessment of parents’ lifestyle (e.g., occupation, smoking and alcohol habits, household cleaning activities, and cooking methods at home) (26), and child’s lifestyle (e.g., physical activity, screen time, diet, cosmetic and hygiene products use, activities at home/school, and drinking water habits) (26,35–39), chronotype (40), smoking and alcohol habits (relevant for children aged 10-11) (26), and exposure directly prior (past 24 or 48 hours) to the sampling (26).

### 3.6 Internal exposome domain

#### 3.6.1 Multi-omics

While other biospecimens, such as hair or nails are not excluded, the main biospecimen will be urine and saliva samples being archived in the Biobank of the University of Cyprus, which is part of the biobank.cy Center of Excellence in Biobanking and Biomedical Research, and in respective biobanks of participating countries. Such samples will be invaluable for multi-omics profiling to explore molecular mechanisms linking exposures with outcomes, capturing both intermediate biological responses and early biological effect biomarkers. Urine samples will be subjected to untargeted GC–MS metabolomics analysis (41). Metabolomics will be used as an intermediate biological layer between biomarkers of exposure and biomarkers of effect. Saliva samples will be used for transcriptomic profiling to detect RNA biomarkers associated with early biological programming and disease susceptibility (42). Extracting RNA from saliva can offer insight into inflammation pathways, which are usually modified before signs of disease appear. Urine proteomics will be used to identify protein biomarkers related to metabolic and systemic functions (43). Proteomic signatures can reflect early-stage biomarkers, thus identifying early deviations from a healthy status.

#### 3.6.2 Oxidative stress/damage and inflammation

Competitive ELISA or mass spectrometry will be used to determine urinary concentrations of oxidative stress/damage and inflammation biomarkers (i.e., 8-iso-PGF2α, 8-OHdG, C-reactive protein, 4-HNE). Creatinine-corrected biomarker values will be calculated after measurements of urinary creatinine using the colorimetric Jaffé method (44).

#### 3.6.3 Iodine, iron deficiencies

Urine samples will be analysed for total iodine following the CDC Environmental Health protocol ITU004A (45) using inductively coupled plasma mass spectrometer (ICP-MS; Thermo X Series II, Thermo Scientific). Based on the WHO classification scheme (46), for school-age children (≥6 years of age), an adequate iodine level is defined as a population median urinary iodine concentration of 100-199 μg/L, whereas a population median of <100 μg/L indicates that the population’s iodine intake is insufficient. Similar analyses for iron and potentially for vitamin D will be followed.

#### 3.6.4 Corticosteroids and circadian rhythm markers

GC-MS methods will be used to determine urinary/salivary concentrations of corticosteroids, including, cortisol, estradiol, testosterone, among others. (47), including melatonin as a marker of circadian rhythm.

#### 3.6.5 Liver and renal function

Biomarkers of liver and renal function that will be measured in urine samples, include glucose, bilirubin, ketones, specific gravity, blood, pH, protein, urobilinogen, nitrite, creatinine, leucocytes, albumin, total protein, and calcium with conventional biochemical spectrophotometry assay.

#### 3.6.6 Intrinsic properties and medical history

Standardized questions based on validated questionnaires will be used for the assessment of each child’s intrinsic characteristics (e.g. age, sex) (26) and medical history (e.g. parents’ diseases, child’s diseases, medicine use, birth anthropometrics, and perinatal medical history) (23,26).

## 4 Main Cohort Outcomes

### 4.1.1 Neurodevelopmental outcomes

The validated screening tool for Social Responsiveness Scale (SRS-1) as a screening tool for autism spectrum disorders (48,49) and the KINDL quality of life questionnaire (50) will be used.

### 4.1.2 Cardio-metabolic outcomes

Waist circumference will be measured, and BMI age-and sex-standardized z-scores will be calculated based on the measurements of weight and height. Systolic and diastolic blood pressure will be measured using automatic upper arm sphygmomanometers.

### 4.1.3 Respiratory Outcomes

The International Study of Asthma and Allergies in Childhood (ISAAC) questionnaires will be used to assess the prevalence and severity of asthma and allergic disease in children (51).

### 4.2 Statistical considerations

#### 4.2.1 Sample size calculation

Estimating the sample size in this type of study (child cohort exposome study) presents several challenges, due to the need to account for multiple factors, including the repeated measures design, the analytical variability, the multiplicity of exposures, the correction for multiple testing, and the minimum detectable concentration difference [59], [60]. A sample size of 300 children per country was calculated to permit detection of standardized differences of approximately 0.3 over six assessment periods, with 80% power, a significance level of 0.05, and 10% annual attrition. This is expected to yield a final analytic sample of approximately 200 children per country, retained over the minimum of 6 years study duration.

#### 4.2.2 Indicative statistical analyses

For the characterization of the spatiotemporal variability of the exposomic profiles, descriptive statistics will be used, including the mean, median, standard deviation, and interquartile range. To visualize the variation in high-dimensional exposome data, principal component analysis (PCA) will be used and to identify patterns in exposure profiles, clustering techniques will be employed (52,53). Furthermore, in order to model children’s exposomic profiling over space and time, linear mixed models (LMMs), generalized linear models, and Bayesian hierarchical models will be applied, as appropriate. For handling missing data, imputation techniques, such as MICE and missForest, will be utilized (54). FDR adjustment will be used, as appropriate, for multiple comparisons correction (55).

In order to estimate the effect of single exposures on early-life biological markers (such as inflammation and oxidative stress), multivariable regression (linear/logistic/mixed) models will be used, adjusting for confounders, such as exposome-wide association models (ExWAS) (56). For the exploration of multiple exposures and outcomes association, variable selection algorithms including deletion-substitution-addition (DSA), elastic net (ENET), Bayesian kernel machine regression (BKMR), lagged kernel machine regression (LKMR), hierarchical clustering on principal component, partial least squares-discriminant analysis (PLS-DA) will be used, as needed (52,57). Weighted quantile sum (WQS) regression will be applied to assess cumulative mixture effects (52). Structural equation modeling (SEM) and causal mediation analysis will be used to explore whether biomarkers of effect mediate the pathway from exposures to health outcomes (58). Latent class analysis (LCA) will be applied to group children into exposure and effect clusters (59). Metabolomics will be used as an intermediate layer between exposures and outcomes with pathway analysis used for transcriptomics and biological enrichment analysis for proteomics.

To generate NCD risk prediction groups, machine learning algorithms, such as random forests, XGBoost, and neural networks will be used, while LASSO will be used for feature selection (52,60). Unsupervised clustering, like k-means, will be used to define NCD risk groups. Targeted maximum likelihood estimation could also be used to estimate causal effects of modifiable exposures on predicted NCD risk groups (61).

#### 4.2.3 Federated data analysis using a Common Data Model

To exploit the multi-country site focus of CHILDREN_FIRST, all data collection methods and study instruments have been *a priori* agreed upon by all participating countries to facilitate federated data processing. To this extent, the reference model of Observational Medical Outcomes Partnership – Common Data Model (OMOP-CDM), as part of the Observational Health Data Sciences and Informatics (OHDSI), will be used (62). OHDSI is an open initiative to facilitate the analysis of real-world data via observational studies. On top of this reference data model, various software tools and methodological approaches have been developed and used extensively (statistical analysis approaches, reference dictionaries) to support the execution of observational studies. The European Health Data & Evidence Network (EHDEN) is a European initiative which aims to implement the OHDSI platform and set up a network of partners in Europe (63). Notably, EHDEN has (so far) attracted more than 180 data partners across Europe, significantly contributing to the setup of the global OHDSI network. Thus, OMOP-CDM has been selected to be used by the CHILDREN_FIRST consortium as it can be considered the de facto standard for real-world data analysis in Europe.

Practically, this means that all CHILDREN_FIRST cohort sites will convert their data to OMOP-CDM. This will allow the formulation of a research question based on the reference CDM, which can then be distributed to each partner site following a “federated” paradigm of data analysis. Each site executes the query locally, and the results are aggregated and analyzed centrally. The advantage of this approach is that there is no need to transfer raw personal/sensitive data, thus avoiding legal and ethical challenges, while robust, large-scale observational analyses across the various populations can take place.

### 4.3 Data management

Data use, similarly to biosample use, is governed by the CHILDREN_FIRST Data Access Committee (DAC) and follows the EU GDPR data sharing policy and governed by sample access requirements for biobanks. A data management plan will be developed following the principles of Open Access by the European Commission. The plan will address the issues of data cleaning, integration, assessment, and sharing including future publications guidelines and it will be aligned with the GDPR 2016/679 regulation for privacy protection and confidentiality. The data management plan, which will be developed during the project’s planning phase, prior to the initiation of data collection, will represent a mutual understanding between all the project collaborators and will be regularly updated as the project evolves.

Standard Operating Procedures (SOPs) for all study-related activities such as recruitment procedures, field and questionnaire data collection and follow-up assessments, will be developed before the study commences. All study instruments (questionnaires, measurement devices, e.g., sensors, sphygmomanometers, scales, measuring tapes) will be piloted prior to the main study initiation. All SOPs will be documented in a General Survey Manual and will be finalized after their testing during the feasibility phase of the study. To ensure compliance with SOPs, training of all involved collaborators will take place.

Compliance with good scientific practice in general and the agreed SOPs in particular will be ensured by regular reporting schemes. Quality control and assurance will encompass documentation and validation of databases as well as software routines for data warehousing and analysis. Training of project team members will take place for different activities of the study: field/data collection activities, effective engagement with participants, and data management.

An electronic toolbox-hub will be developed following the establishment of the study, and it will comprise various interfaces tailored to different user groups: (1) Internal: Research team members will have password-protected access to study data and metadata. All data will be de-identified and pseudo-anonymized to ensure participants’ confidentiality. (2) Parental: Parents will have password-protected access to their child’s individual health and other data. This interface will include personalized reports on biomarker levels accompanied by interpretative information, such as reference ranges (where available), comparisons with age-matched population averages, and relevant recommendations. (3) Aggregated: Policy makers and school authorities will have access to aggregated data (with the city as the unit of reference). This will include summaries of various biomarker levels and health-related outcomes measured across the study population. (4) Public: The public will have access to scientific findings in the form of flyers and infographics. (5) Open access: Datasets will be available upon request after the submission and evaluation of a data application by interested research teams.

### 4.4 Ethics and dissemination

The study protocol has been approved by the Cyprus National Bioethics Committee (EEBK/EP/2022/68) and the Cyprus Ministry of Education, Sports and Youth (07.15.005.011.001).

All study procedures will be conducted in accordance with the principles of the World Medical Association Declaration of Helsinki, and the study will have local ethics approval, abiding by the ethical standards of each participating country. Written informed consent will be available in each country’s official language and in English, and it will be obtained from parents/guardians. Verbal assent will be required by each child before any type of assessment. Parents have the right to withdraw approval for their child’s participation at any stage, for any reason without having to give any explanations and without any consequences for the participant. The study does not involve any risks, since the sampling is non-invasive. All data will be treated with strict confidentiality. A unique identification code will be assigned to each participating child and will be used in questionnaires, biological samples, and databases.

Study results will be disseminated at regional and international conferences and in peer-reviewed journals. Different types of results (personalized, aggregated) will be available to distinct user groups, i.e., parents and schools, respectively, through the electronic toolbox-hub (see the Data management section). Aggregated cohort-level results will be communicated annually through newsletters. Until the electronic toolbox-hub is fully developed, individualized reports will be delivered to parents on an annual basis through a secure, password-protected system. These reports will include detailed interpretations of the assessed parameters and personalized recommendations tailored to the child’s data.

### 4.5 Patient and public involvement statement

An integral part for the success of CHILDREN_FIRST is the active engagement of participants, their families, and the school personnel. To facilitate this, a multifaceted engagement approach will be applied. Prior to study initiation, focus groups with parents, teachers, and headmasters will be conducted to discuss the study instruments, the ethical and security issues including the risks and benefits of the CHILDREN FIRST study protocol and gather their perspectives on strategies for maintaining participation throughout follow-up assessments. A feasibility study with a small number of participants will take place to assess response rates and key study procedures, including recruitment, data collection, and the transfer and storage of samples.

An Advisory Board, comprising experts in environmental epidemiology, biomedical sciences, exposure science, the exposome field, and the entrepreneurship sector will be established to support the study. The Advisory Board will play a critical role in monitoring and providing constructive input on the study’s progress, offering recommendations for addressing emerging challenges, advising on strategic planning, and contributing insights to enhance the study’s societal impact.

A Participant Advisory Committee, composed of interested parents of children participating in the study, will be assembled to provide ongoing input. The committee will meet regularly to discuss various topics, including data collection procedures, reports of individualized results, and the incorporation of participant feedback.

Research staff will be trained to have an effective dialogue with children and their parents/guardians so that they can help promote participants’ empowerment and ensure long-term participant engagement. Regular communication will be maintained through personal interactions, phone calls and emails. A dedicated hotline at each site will be available for parents to ask questions or share concerns.

The expected benefit for participants is that they will receive personalized reports containing selected data related to their children’s assessments. Moreover, periodic newsletters will be distributed, providing updates on the study’s progress and offering opportunities for feedback. All children will also receive a symbolic gift voucher for each annual assessment.

## 5 Discussion

CHILDREN_FIRST is unique in its scope and design, representing the first *a priori* harmonized, exposome-based children’s cohort that will conduct annual follow-ups across five Mediterranean countries during the critical windows of vulnerability (6-11 years of age). The CHILDREN_FIRST study integrates the exposome concept and personalized prevention approaches to examine multiple environmental exposures and health outcomes over time. The anticipated impact can be outlined under two key pillars: Methodological novelties and generation of novel knowledge.

### 5.1 Methodological novelties

#### 5.1.1 First exposomics-based children’s prospective cohort in the Mediterranean region

CHILDREN_FIRST will be the first exposomics-based children’s longitudinal study in the Mediterranean countries of Cyprus, Greece, Spain, Albania, and Israel. Unlike most pregnancy-birth cohorts, CHILDREN_FIRST targets ages 6 to <11 years upon child’s entry to primary school, which is a relatively understudied, yet critical developmental period of life (8). This study will conduct annual assessments beginning in first grade all the way through the final year of primary school, collecting a maximum of six repeated measurements per child over six years. The integration of multi-omics platform will be key in disentangling the gene expression patterns and the downstream biological processes that govern children’s growth and development trajectories in the co-occurrence of a suite of environmental stressors.

#### 5.1.2 Development of precision prevention-based datasets for informed policy making and development of health interventions

The collection of annual cohort data and the generation of NCD-stratified disease risk groups by their exposomic profiles during a critical window of susceptibility will generate robust scientific evidence to support the development of evidence-based guidelines and interventions. These will be disseminated to relevant policymakers, including the Ministries of Health and Education in all participating countries, the WHO/UNESCO/UNICEF health-promoting schools Unit, and beyond. By implementing such measures, the study aims to contribute to reducing the future incidence of NCDs, hence lowering governmental healthcare expenditures and broader societal costs.

#### 5.1.3 Longer-term cohort infrastructure for future exposomic research and collaboration

The CHILDREN_FIRST cohort will produce spatio-temporal data on child health of high-quality, complemented by biospecimen collection and storage to biobanks and an electronic toolbox-hub. This infrastructure will not only support the current study objectives but will also serve as a long-term resource for hypothesis generation and testing and it will facilitate collaboration with local and EU institutions as well as with other cohort studies.

#### 5.1.4 Emphasis on the school setting

The CHILDREN_FIRST cohort has intentionally focused on the school setting to inform, recruit, collect data and samples, and inform parents and many stakeholders as part of the cohort’s related activities. This is a cost-efficient approach to engaging stakeholders in the children’s health prevention programs towards achieving healthy growth and development for all children, leaving no one behind.

### 5.2 Generation of novel knowledge

#### 5.2.1 Understanding environmental impacts on early disease development

CHILDREN_FIRST aims to advance the understanding of how multiple environmental exposures affect the development of diseases and the progression of early-stage disease markers during the vulnerable developmental window of ages 6 to 11 years.

#### 5.2.2 Uncovering the temporal biodynamics of early-stage disease markers

The collection of a high number of repeated measurements during a critical life stage will allow detailed investigation of the temporal dynamics of disease processes. Particular emphasis will be given to early-stage disease markers, such as biomarkers of oxidative stress, inflammation, and omics-based signatures.

While changes in anthropometric indicators, such as BMI, are expected within one-year intervals, data on the temporal trend of obesogenic molecular markers in this age group remain scarce. The CHILDREN_FIRST study will be instrumental in identifying key windows of susceptibility and the timing of potential interventions.

For example, existing evidence suggests that interventions as early as at 6 months old can significantly improve developmental trajectories in children at risk for autism spectrum disorders (64). Similarly, global data indicate that the prevalence of overweight among children aged 5–19 years (18% in 2016) is considerably higher than in children under five (5.6% in 2019) (65). Assessing how BMI and related biomarkers evolve during primary school years could provide critical insights into the metabolic disease processes. Similar hypotheses could also extend to other outcomes, such as cardiometabolic and neurodevelopmental.

#### 5.2.3 Identification of novel early-stage biomarkers

The application of multiple omics platforms, such as metabolomics, will facilitate the comprehensive profiling of the children’s exposome. Multi-omics platforms, such as genomics, transcriptomics, epigenomics, microbiomics, proteomics and metabolomics may be used as intermediaries, i.e., mediators between main exposomic variables and the key outcomes or end points of disease. Machine learning algorithms will explore associations between omics signatures and both exposure profiles and health outcomes, potentially leading to the identification of novel biomarkers that serve as early indicators of chronic disease risk or lead to NCD risk prediction groups with distinct exposomic profiles.

### 5.3 What does this prospective children’s exposomics cohort study add?

CHILDREN_FIRST introduces several innovative contributions to biomedical and environmental health research for children, particularly within the context of the broader Mediterranean region and its heterogeneous populations:

#### 5.3.1 Prospective health data collection for middle childhood

The cohort will collect longitudinal population-level health data specifically for children aged 6–11 years in Cyprus, Greece, Spain, Albania, and Israel.

#### 5.3.2 Spatio-temporal characterization of the child exposome

The study will generate novel insights into the spatial and temporal variability of exposome data within the child population in these five countries, improving the understanding of how environmental exposures differ across time and geographical area.

#### 5.3.3 High-frequency, repeated exposomic measurements with non-invasive biomarkers of exposure/effect

By conducting six assessment points over six consecutive years, this study enables repeated data collection within a relatively short timeframe, an approach that enhances the ability to detect temporal trends and exposomic trajectories that would prospectively frame up children’s growth and development curves.

#### 5.3.4 Assessment of novel environmental stressors

The cohort includes evaluation of emerging and underexplored exposures, such as artificial light at night (ALAN) or microplastics or PFAS, among others, and investigates their potential health impact on children’s growth and development.

#### 5.3.5 Integration of environmental and health profiles using exposomics

CHILDREN_FIRST will apply an exposomic framework to integrate environmental exposure data with health outcomes, leveraging advanced tools for comprehensive profiling of the environment–health interface.

#### 5.3.6 Stakeholder-specific data dissemination via an electronic toolbox-hub

A secure digital platform (toolbox-hub) will facilitate access to study findings, with tailored interfaces for different stakeholders: (1) Parents will receive personalized reports for their child, including biomarker interpretations and recommendations. (2) Policymakers and school authorities will be able to access aggregated, city-level data to inform public health strategies. (3) Researchers will have access to de-identified datasets for further analysis.

#### 5.3.7 Adoption of common data models

Multi-country cohort data will be processed using the OMOP-CDM, promoting data interoperability, facilitating collaboration and enhancing the potential for data reuse in future research, including secondary use in future studies.

### 5.4 Limitations and Strengths

Unlike most birth cohorts that collect data during pregnancy, our study begins recruitment at the start of primary school. However, data on birth and perinatal periods will be retrospectively gathered through validated questionnaires completed by parents, which may introduce recall bias. The sample size may be relatively modest (n=200-300 per site), but the longitudinal design—featuring six repeated measurements over six years—will yield approximately 1200-1800 data points per cohort site, providing rich datasets with enhanced statistical power. On the other hand, CHILDREN_FIRST will be the first longitudinal, exposome-based and a priori harmonized children’s multi-country site cohort study with annual follow-ups in five Mediterranean countries — Cyprus, Greece, Spain, Albania, and Israel — using the same study instruments/tools. Study procedures, such as recruitment, assessments and reporting back, will be centered on the school setting, transforming schools into health-promoting schools (HPS), based on the WHO/UNESCO/UNICEF HPS paradigm towards integrating health into all aspects of school life. The high number of repeated measurements during a pre-defined critical life stage of vulnerability (6-11 years), with baseline at entry in primary school (around 6 years of age) will allow for delineating the temporal dynamics of chronic disease processes as impacted by children’s exposomic trajectories during this critical developmental window. The spatiotemporal profiling of children’s exposomes will provide the basis for exploration of associations between multi-omics signatures, biomarkers of exposure/effect profiles and health outcomes, potentially identifying novel non-genetic markers as early indicators of chronic disease risk. The collection of annual cohort data and the generation of NCD-stratified disease risk groups by their exposomic profiles during a critical window of susceptibility will generate robust scientific data to inform evidence-based guidelines and non-pharmacological health interventions.

### 5.5 Future directions and implications

CHILDREN_FIRST is an ambitious initiative that will establish a long-term prospective cohort infrastructure for biomedical research on children’s health within the Mediterranean region linked with established biobanks, such as the biobank.cy. It will be key in framing and advancing the epidemiological surveillance of chronic diseases during childhood, in collaboration with relevant institutions, public health bodies and universities. This study is aligned with the European Commission’s principles of personalized medicine. The cohort findings will systematically feed into the evaluation and design of chronic disease prevention programs. Expected results would inform evidence-based policy making and the development of health interventions for reducing the risk of NCDs. The effective implementation of such disease prevention and control programs fed by CHILDREN_FIRST cohort results would generate substantial reductions in social costs of NCDs with longer-term benefits for the health sector, the quality of life, and wellbeing.

## Data Availability

No datasets were generated or analysed during the current study. All relevant data from this study will be made available upon study completion.

## 6 Acknowledgments

We would like to express our sincere gratitude to the Cyprus Ministry of Education, Youth and Sports and to the Ministry of Health in Cyprus for their strong support. We are also deeply grateful to the Cyprus Pediatric Society for their support and their invaluable guidance.

## 7 Supporting information

Supplementary File 1: Methodological details

Supplementary File 2: SPIROS checklist

## 8 Funding acquisition

This research received start-up funds from internal funds and donors in Cyprus. ISGlobal acknowledge support from the grant CEX2023-0001290-S funded by MCIN/AEI/10.13039/501100011033, and support from the Generalitat de Catalunya through the CERCA Program. The biobank.cy Center of Excellence which hosts the Biobank of Cyprus is funded through the *CY*-Biobank, which is an EU Horizon 2020 Research and Innovation Programme under Grant Agreement no. 857122, the Republic of Cyprus, and the University of Cyprus.

## 9 Author Contributions

KCM: conceptualisation, supervision, methodology, resources, project administration, funding and writing–original draft and review-editing. CK: conceptualisation, methodology, project administration and writing–original draft and review-editing. GS: methodology and data curation, writing–review and editing. CD: supervision, methodology, resources, project administration, funding and writing–review-editing. CAC: methodology and writing–review-editing. MG: methodology and writing–review-editing. EAM: methodology and writing– review-editing. IG: methodology clinical data curation, and writing–review-editing. NB: methodology, supervision, and writing–review-editing. IM: methodology and writing–review-editing. OD: methodology and writing–review-editing. PN: methodology and writing–review-editing. ZT: methodology and writing–review-editing. LE: methodology and writing–review-editing. AK: clinical data curation, methodology and writing–review-editing. EAE: methodology and writing–review-editing. PC: methodology and writing–review-editing. AA: methodology and writing–review-editing. GM: methodology and writing–review-editing. CH: clinical data curation, methodology and writing–review-editing. MT: methodology, resources, supervision, project administration and writing–review-editing. TM: methodology and writing–review-editing. AK: methodology and writing–review-editing. All authors approved the publication of the protocol.

## References

1. Bloom DE, Cafiero ET, Jané-Llopis E, Abrahams-Gessel S, Bloom LR, Fathima S, et al. The Global Economic Burden of Noncommunicable Diseases. Geneva: World Economic Forum; 2011.

2. Armocida B, Monasta L, Sawyer S, Bustreo F, Segafredo G, Castelpietra G, et al. Burden of non-communicable diseases among adolescents aged 10–24 years in the EU, 1990–2019: a systematic analysis of the Global Burden of Diseases Study 2019. Lancet Child Adolesc Health. 2022 Jun;6(6):367–83.

3. GBD 2019 Cancer Risk Factors Collaborators. The global burden of cancer attributable to risk factors, 2010-19: a systematic analysis for the Global Burden of Disease Study 2019. Lancet Lond Engl. 2022 Aug 20;400(10352):563–91.

4. World Health Organization. Global action plan for the prevention and control of noncommunicable diseases 2013-2020 [Internet]. Geneva: World Health Organization; 2013 [cited 2022 Jul 7]. Available from: https://apps.who.int/iris/handle/10665/94384

5. UN General Assembly. Transforming our world: the 2030 Agenda for Sustainable Development [Internet]. 2015. Report No.: A/RES/70/1. Available from: https://sdgs.un.org/sites/default/files/publications/21252030%20Agenda%20for%20Sustainable%20Development%20web.pdf

6. World Health Organization. Thirteenth General Programme of Work, 2019–2023 [Internet]. Geneva: World Health Organization; 2018. Available from: http://www.who.int/about/what-wedo/gpw-thirteen-consultation/en/

7. Landrigan PJ, Kimmel CA, Correa A, Eskenazi B. Children’s health and the environment: public health issues and challenges for risk assessment. Environ Health Perspect. 2004 Feb;112(2):257–65.

8. Cohen Hubal EA, de Wet T, Du Toit L, Firestone MP, Ruchirawat M, van Engelen J, et al. Identifying important life stages for monitoring and assessing risks from exposures to environmental contaminants: Results of a World Health Organization review. Regul Toxicol Pharmacol. 2014 Jun;69(1):113–24.

9. Ward ZJ, Long MW, Resch SC, Giles CM, Cradock AL, Gortmaker SL. Simulation of Growth Trajectories of Childhood Obesity into Adulthood. N Engl J Med. 2017 Nov 30;377(22):2145–53.

10. Bui DS, Lodge CJ, Burgess JA, Lowe AJ, Perret J, Bui MQ, et al. Childhood predictors of lung function trajectories and future COPD risk: a prospective cohort study from the first to the sixth decade of life. Lancet Respir Med. 2018 Jul;6(7):535–44.

11. European Parliament. Directorate General for Parliamentary Research Services. Human exposome research: potentials, limitations and public policy implications. [Internet]. LU: Publications Office; 2025 [cited 2025 May 26]. Available from: https://data.europa.eu/doi/10.2861/5600247

12. Wild CP. Complementing the genome with an ‘exposome’: the outstanding challenge of environmental exposure measurement in molecular epidemiology. Vol. 14, Cancer epidemiology, biomarkers & prevention: a publication of the American Association for Cancer Research, cosponsored by the American Society of Preventive Oncology. 2005. p. 1847–50.

13. Wild CP. The exposome: from concept to utility. Int J Epidemiol. 2012 Feb 1;41(1):24–32.

14. Haddad N, Andrianou XD, Makris KC. A Scoping Review on the Characteristics of Human Exposome Studies. Curr Pollut Rep. 2019;5(4):378–93.

15. Willett WC. Balancing Life-Style and Genomics Research for Disease Prevention. Science. 2002 Apr 26;296(5568):695–8.

16. European Commission. Personalised Medicine [Internet]. 2020. Available from: https://research-and-innovation.ec.europa.eu/research-area/health/personalised-medicine_en

17. Gambhir SS, Ge TJ, Vermesh O, Spitler R. Toward achieving precision health. Sci Transl Med. 2018 Feb 28;10(430):eaao3612.

18. Vrijheid M, Slama R, Robinson O, Chatzi L, Coen M, Van Den Hazel P, et al. The Human Early-Life Exposome (HELIX): Project Rationale and Design. Environ Health Perspect. 2014 Jun;122(6):535–44.

19. Maitre L, De Bont J, Casas M, Robinson O, Aasvang GM, Agier L, et al. Human Early Life Exposome (HELIX) study: a European population-based exposome cohort. BMJ Open. 2018 Sep;8(9):e021311.

20. Jacobson LP, Parker CB, Cella D, Mroczek DK, Lester BM, on behalf of program collaborators for Environmental influences on Child Health Outcomes, et al. Approaches to protocol standardization and data harmonization in the ECHO-wide cohort study. Pediatr Res. 2024 Jun;95(7):1726–33.

21. Hughes RA, Tilling K, Lawlor DA. Combining Longitudinal Data From Different Cohorts to Examine the Life-Course Trajectory. Am J Epidemiol. 2021 Dec 1;190(12):2680–9.

22. Wild CP. The exposome: from concept to utility. Int J Epidemiol. 2012 Feb;41(1):24–32.

23. EURO-URHIS 2. European Urban Health Indicators Part Two: Using indicators to inform policy [Internet]. 2014 Dec. Available from: https://cordis.europa.eu/project/id/223711/reporting

24. Statistical Service of Cyprus. European Health Interview Survey [Internet]. 2016 Nov. Available from: https://library.cystat.gov.cy/Documents/Questionnaires/EUROPEAN_HEALTH_SURVEY-2014-EN.pdf

25. Statistical Service of Cyprus. Survey on Income and Living Conditions of Households 2022 [Internet]. 2022. Available from: https://www.cystat.gov.cy/en/QuestionnaireList?s=44

26. González-Alzaga B, Hernández AF, Kim Pack L, Iavicoli I, Tolonen H, Santonen T, et al. The questionnaire design process in the European Human Biomonitoring Initiative (HBM4EU). Environ Int. 2022 Feb;160:107071.

27. Charisiadis P, Andra SS, Makris KC, Christophi CA, Skarlatos D, Vamvakousis V, et al. Spatial and seasonal variability of tap water disinfection by-products within distribution pipe networks. Sci Total Environ. 2015 Feb;506–507:26–35.

28. Nomura H, Ueyama J, Kondo T, Saito I, Murata K, Iwata T, et al. Quantitation of neonicotinoid metabolites in human urine using GC-MS. J Chromatogr B Analyt Technol Biomed Life Sci. 2013 Dec;941:109–15.

29. Leng G, Gries W. Determination of Pyrethroids in Blood Plasma and Pyrethroid/Pyrethrin Metabolites in Urine by Gas Chromatography-Mass Spectrometry and High-Resolution GC-MS BT - Pesticide Protocols. In: Martínez Vidal JL, Frenich AG, editors. Totowa, NJ: Humana Press; 2006. p. 17–33. Available from: 10.1385/1-59259-929-X:017

30. CDC. Laboratory Procedure Manual. Matrix: Urine. Method: Urine Multi-Element ICP-DRC-MS Renamed from Inductively Coupled Plasma-Mass Spectrometry (ICP-DRC-MS). Method No: 3018.3 (15 element panel) and 3018A.2 (total arsenic). [Internet]. Centers for Disease Control and Prevention. Inorganic Radionuclides and Toxicology, Division of Laboratory Sciences, National Center for Environmental Health; 2012 p. 1–48. Available from: https://www.cdc.gov/nchs/data/nhanes/nhanes_11_12/uhm_g_met_heavy_metals.pdf

31. Kalyvas H, Andra SS, Charisiadis P, Karaolis C, Makris KC. Influence of household cleaning practices on the magnitude and variability of urinary monochlorinated bisphenol A. Sci Total Environ. 2014;490:254–61.

32. ISGlobal. HELIX Project: Data Inventory (Subcohort) [Internet]. 2014 [cited 2023 Jan 20]. Available from: http://www.projecthelix.eu/data-inventory

33. Baloch RM, Maesano CN, Christoffersen J, Banerjee S, Gabriel M, Csobod É, et al. Indoor air pollution, physical and comfort parameters related to schoolchildren’s health: Data from the European SINPHONIE study. Sci Total Environ. 2020;739:139870.

34. Garcia-Saenz A, Sánchez de Miguel A, Espinosa A, Valentin A, Aragonés N, Llorca J, et al. Evaluating the Association between Artificial Light-at-Night Exposure and Breast and Prostate Cancer Risk in Spain (MCC-Spain Study). Environ Health Perspect. 2018 Apr 5;126(4):047011.

35. Makris KC, Konstantinou C, Andrianou XD, Charisiadis P, Kyriacou A, Gribble MO, et al. A cluster-randomized crossover trial of organic diet impact on biomarkers of exposure to pesticides and biomarkers of oxidative stress/inflammation in primary school children. PLoS ONE. 2019;14(9):1–15.

36. Klakk H, Wester CT, Olesen LG, Rasmussen MG, Kristensen PL, Pedersen J, et al. The development of a questionnaire to assess leisure time screen-based media use and its proximal correlates in children (SCREENS-Q). BMC Public Health. 2020 Dec;20(1):664.

37. López-Gajardo MA, Leo FM, Sánchez-Miguel PA, López-Gajardo D, Soulas C, Tapia-Serrano MA. KIDMED 2.0, An update of the KIDMED questionnaire: Evaluation of the psychometric properties in youth. Front Nutr. 2022 Nov 8;9:945721.

38. Bruni O, Ottaviano S, Guidetti V, Romoli M, Innocenzi M, Cortesi F, et al. The Sleep Disturbance Scale for Children (SDSC) Construct ion and validation of an instrument to evaluate sleep disturbances in childhood and adolescence. J Sleep Res. 1996 Dec;5(4):251–61.

39. Foraster M, Künzli N, Aguilera I, Rivera M, Agis D, Vila J, et al. High Blood Pressure and Long-Term Exposure to Indoor Noise and Air Pollution from Road Traffic. Environ Health Perspect. 2014 Nov;122(11):1193–200.

40. Werner H, LeBourgeois MK, Geiger A, Jenni OG. Assessment of Chronotype in Four- to Eleven-Year-Old Children: Reliability and Validity of the Children’s ChronoType Questionnaire (CCTQ). Chronobiol Int. 2009 Jan;26(5):992–1014.

41. Chan ECY, Pasikanti KK, Nicholson JK. Global urinary metabolic profiling procedures using gas chromatography-mass spectrometry. Nat Protoc. 2011;6(10):1483–99.

42. Palanisamy V, Wong DT. Transcriptomic Analyses of Saliva. In: Seymour GJ, Cullinan MP, Heng NCK, editors. Oral Biology [Internet]. Totowa, NJ: Humana Press; 2010 [cited 2025 Apr 29]. p. 43–51. (Methods in Molecular Biology; vol. 666). Available from: http://link.springer.com/10.1007/978-1-60761-820-1_4

43. Joshi N, Garapati K, Ghose V, Kandasamy RK, Pandey A. Recent progress in mass spectrometry-based urinary proteomics. Clin Proteomics. 2024 Dec;21(1):14.

44. Angerer J, editors Hartwig A. The MAK-Collection for Occupational Health and Safety. In: Angerer J, Hartwig A, editors. The MAK-Collection for Occupational Health and Safety. Weinheim: Wiley-VCH Verlag GmbH & Co. KGaA; 2010. p. 169–84.

45. CDC. Urine iodine ICPMS [Renamed from inductively coupled plasma-mass spectrometry (ICP-MS)], ITU004A [Internet]. 2007 Dec. Available from: https://www.cdc.gov/nchs/data/nhanes/nhanes_03_04/l06uio_c_met_urine_iodine_ICPMS.pdf

46. WHO. Iodine deficiency [Internet]. 2013. Available from: https://www.who.int/data/nutrition/nlis/info/iodine-deficiency

47. Moon JY, Jung HJ, Moon MH, Chung BC, Choi MH. Heat-map visualization of gas chromatography-mass spectrometry based quantitative signatures on steroid metabolism. J Am Soc Mass Spectrom. 2009 Sep 1;20(9):1626–37.

48. Constantino JN. The Social Responsiveness Scale. Los Angeles: Los Angeles: Western Psychological Services; 2002.

49. Constantino JN, Gruber CP. Social Responsiveness Scale: Manual. Los Angeles: Western Psychological Services; 2005.

50. Ravens-Sieberer U, Bullinger M. Assessing health-related quality of life in chronically ill children with the German KINDL: first psychometric and content analytical results. Qual Life Res. 1998 Jul;7(5):399–407.

51. Asher, Weiland. The International Study of Asthma and Allergies in Childhood (ISAAC). Clin Exp Allergy. 1998 Nov;28(s5):52–66.

52. Maitre L, Guimbaud JB, Warembourg C, Güil-Oumrait N, Petrone PM, Chadeau-Hyam M, et al. State-of-the-art methods for exposure-health studies: Results from the exposome data challenge event. Environ Int. 2022 Oct;168:107422.

53. Chang L, Ewald J, Hui F, Bayen S, Xia J. A data-centric perspective on exposomics data analysis. Exposome [Internet]. 2024 Feb 9 [cited 2025 May 23];4(1). Available from: https://academic.oup.com/exposome/article/doi/10.1093/exposome/osae005/7657695

54. Sun Y, Li J, Xu Y, Zhang T, Wang X. Deep learning versus conventional methods for missing data imputation: A review and comparative study. Expert Syst Appl. 2023 Oct;227:120201.

55. Chen SY, Feng Z, Yi X. A general introduction to adjustment for multiple comparisons. J Thorac Dis. 2017 Jun;9(6):1725–9.

56. Chung MK, House JS, Akhtari FS, Makris KC, Langston MA, Islam KT, et al. Decoding the exposome: data science methodologies and implications in exposome-wide association studies (ExWASs). Exposome [Internet]. 2024 Feb 9 [cited 2025 May 23];4(1). Available from: https://academic.oup.com/exposome/article/doi/10.1093/exposome/osae001/7574628

57. Warembourg C, Anguita-Ruiz A, Siroux V, Slama R, Vrijheid M, Richiardi L, et al. Statistical Approaches to Study Exposome-Health Associations in the Context of Repeated Exposure Data: A Simulation Study. Environ Sci Technol. 2023 Oct 31;57(43):16232–43.

58. Liu SH, Weber ES, Manz KE, McCarthy KJ, Chen Y, Schüffler PJ, et al. Assessing the Impact and Cost-Effectiveness of Exposome Interventions on Alzheimer’s Disease: A Review of Agent-Based Modeling and Other Data Science Methods for Causal Inference. Genes. 2024 Nov 12;15(11):1457.

59. Santaolalla A, Garmo H, Grigoriadis A, Ghuman S, Hammar N, Jungner I, et al. Metabolic profiles to predict long-term cancer and mortality: the use of latent class analysis. BMC Mol Cell Biol [Internet]. 2019 Dec [cited 2025 May 23];20(1). Available from: https://bmcmolcellbiol.biomedcentral.com/articles/10.1186/s12860-019-0210-7

60. Isola S, Murdaca G, Brunetto S, Zumbo E, Tonacci A, Gangemi S. The Use of Artificial Intelligence to Analyze the Exposome in the Development of Chronic Diseases: A Review of the Current Literature. Informatics. 2024 Nov 12;11(4):86.

61. Smith MJ, Phillips RV, Luque-Fernandez MA, Maringe C. Application of targeted maximum likelihood estimation in public health and epidemiological studies: a systematic review. Ann Epidemiol. 2023 Oct;86:34–48.e28.

62. Observational Health Data Sciences and Informatics [Internet]. 2025. Available from: https://www.ohdsi.org/

63. European Health Data & Evidence Network [Internet]. 2025. Available from: https://www.ehden.eu/

64. Bradshaw J, Steiner AM, Gengoux G, Koegel LK. Feasibility and effectiveness of very early intervention for infants at-risk for autism spectrum disorder: a systematic review. J Autism Dev Disord. 2015 Mar;45(3):778–94.

65. World Health Organization. Obesity and overweight [Internet]. 2021 [cited 2022 Jun 27]. Available from: https://www.who.int/news-room/fact-sheets/detail/obesity-and-overweight

